# Rare-variant burden across lysosomal genes implicates sialylation and ganglioside metabolism in Parkinson’s disease

**DOI:** 10.64898/2026.02.18.26346391

**Authors:** Konstantin Senkevich, Sitki Cem Parlar, Cloe Chantereault, Lang Liu, Eric Yu, Uladzislau Rudakou, Jamil Ahmad, Jennifer A. Ruskey, Farnaz Asayesh, Dan Spiegelman, Cheryl Waters, Oury Monchi, Yves Dauvilliers, Nicolas Dupré, Lior Greenbaum, Sharon Hassin-Baer, Irina Miliukhina, Alla Timofeeva, Anton Emelyanov, Sofya Pchelina, Roy N. Alcalay, Ziv Gan-Or

## Abstract

Lysosomal dysfunction is central to Parkinson’s disease pathogenesis, with *GBA1* as the strongest established genetic risk factor. Numerous other genes involved in lysosomal sphingolipid, glycosphingolipid and ceramide metabolism have been proposed as contributors to Parkinson’s disease, underscoring the need for comprehensive genetic analyses across these pathways. We analysed rare variants (minor allele frequency < 0.01) across 36 lysosomal genes (excluding *GBA1*) in 8,267 individuals with Parkinson’s disease and 68,208 controls, including a subset of 793 early-onset Parkinson’s disease (≤50 years) cases. Targeted sequencing was performed in four cohorts at McGill University (3,456 Parkinson’s disease patients and 2,664 controls) and results were combined with whole-genome sequencing data from the UK Biobank (2,848 cases, 62,451 controls), and from the Accelerating Medicines Partnership - Parkinson’s Disease (1,963 cases, 3,093 controls). We analysed the association of rare variants in these genes with Parkinson’s disease using Sequence Kernel Association Test-Optimal (SKAT-O) across variant classes (all rare variants, nonsynonymous, loss-of-function and predicted damaging variants with a Combined Annotation Dependent Depletion (CADD) score >20), with meta-analysis across cohorts. We additionally performed per-domain analyses for variants in gene segments encoding functional domains. False discovery rate correction was applied. Meta-analysis identified a significant association between rare variants in *ST3GAL3* and Parkinson’s disease (Pfdr=0.04). Several additional lysosomal genes showed nominal associations (P<0.05), including *HGSNAT, ASAH1, CTSD, HEXA, ST3GAL4* and *SGPP1*. Domain-based analyses identified a strong enrichment of nonsynonymous variants within the beta-acetyl-hexosaminidase–like domain of HEXA (P = 8.0 × 10), although this signal did not survive correction for multiple testing (Pfdr=0.154). In early-onset Parkinson’s disease, domain-based analyses revealed significant associations in *NAGLU* (Pfdr=7.3×10) and *ST3GAL5* (Pfdr=0.03). Together, these results provide genetic evidence that rare variants across multiple lysosomal pathways, particularly those related to sialylation, ganglioside metabolism, ceramide biology, and lysosomal proteolysis, may contribute to Parkinson’s disease susceptibility beyond *GBA1*, highlighting biologically coherent pathways for future replication and functional investigation.

## Introduction

Lysosomal dysfunction is increasingly recognized as a key component of Parkinson’s disease pathogenesis ^1^. Mutations in the *GBA1* gene, encoding beta-glucocerebrosidase, are the most common genetic risk factor for Parkinson’s disease, and both heterozygous and biallelic variants are associated with increased disease risk and rapid progression ^2–4^. Beyond *GBA1*, rare and common variants in other lysosomal genes, such as *GALC, ARSA, CTSB*, *SMPD1, TMEM175, SPTSSB*, *ASAH1*, have also been implicated in Parkinson’s disease susceptibility ^5–9^. Studies have demonstrated that the cumulative effect of multiple risk alleles across the lysosomal pathway influences Parkinson’s disease risk and may also influence neuropathological severity, independent of *GBA1* status ^8,10–12^.

Lysosomal impairment disrupts the autophagy-lysosomal and ubiquitin-proteasome systems. This leads to defective clearance of alpha-synuclein and other substrates and organelles, and promotes aggregation, neuroinflammation and dopaminergic neurodegeneration ^13–15^. *GBA1* mutations exemplify this mechanism, whereby reduced beta-glucocerebrosidase activity leads to increased glucosylceramide and glucosylsphingosine, alpha-synuclein accumulation and lysosomal dysfunction ^16,17^. Consistent with this model, multiple additional lysosomal genes have been shown to modulate alpha-synuclein toxicity and autophagic-lysosomal pathway integrity ^7,18^.

Despite these advances, many other genes involved in the lysosomal sphingolipid and ceramide metabolism pathways could be potential contributors to Parkinson’s disease, emphasizing the need for comprehensive genetic analysis within these pathways. To address this gap, we analysed the burden of rare variants across 36 lysosomal genes in a cohort of 8,267 patients and 68,208 controls, aiming to define the contribution of individual genes and the aggregate lysosomal genetic burden to Parkinson’s disease susceptibility and heterogeneity.

## Materials and methods

### Study population

We analysed 8,267 individuals with Parkinson’s disease and 68,208 controls **(Table 1)**. The studied population included: (i) four cohorts sequenced at McGill University, comprising participants recruited in Quebec, Canada and Montpellier, France ^19^, Columbia University, New York, USA ^20^, Sheba Medical Center, Israel ^21^ and the Pavlov First State Medical University and the Institute of Human Brain, Saint-Petersburg, Russia ^6^ (3,456 cases; 2,664 controls); (ii) the UK Biobank (2,848 cases; 62,451 controls); and (iii) the Accelerating Medicines Partnership - Parkinson’s Disease (AMP-PD) whole-genome sequencing cohorts (1,963 cases; 3,093 controls). A subset of 793 patients with early-onset Parkinson’s disease, defined as an age at onset of ≤ 50 years, was analysed separately given the known association of *GBA1* variants with earlier disease onset ^2^. Diagnoses were established by movement-disorder specialists according to the UK Brain Bank ^22^ or MDS clinical diagnostic criteria ^23^. All participants provided written informed consent under protocols approved by their respective research ethics boards. This research was approved by McGill ethics boards.

**Table 1.** Study population.

| Cohort | All Parkinson's disease cases |  |  | Controls |  |  |
| --- | --- | --- | --- | --- | --- | --- |
|  | N | Mean age (SD) | Males, % | N | Mean age (SD) | Males, % |
| McGill cohort | 1123 | 59.84 (11.09) | 715 (63.7%) | 1196 | 48.43 (15.26) | 541 (45.2%) |
| Sheba cohort | 702 | 60.93 (11.79) | 437 (62.3%) | 551 | 33.22 (7.84) | 316 (57.4%) |
| Columbia cohort | 1122 | 59.82 (11.36) | 736 (65.6%) | 500 | 64.29 (10.02) | 177 (35.4%) |
| Pavlov and Human brain institute cohort | 509 | 66.15 (9.29) | 206 (40.5%) | 417 | 73.61 (10.49) | 172 (41.2%) |
| UKBB | 2,848 | 63.00 (5.25) | 1,065 (37.4%) | 62,451 | 56.79 (8.02) | 33,527 (53.7%) |
| AMP-PD | 1,963 | 64.54 (9.52) | 1,260 (64.2%) | 3,093 | 69.76 (13.02) | 1,496 (48.4%) |
| All cohorts | 8,267 | - |  | 68,208 |  |  |
| Cohort | Parkinson's disease with early onset $\leq 50$ years | | | Controls | | |
| | N | Mean age ± SD | Males, % | N | Mean age ± SD | Males, % |
| McGill cohort | 201 | 44.09 (6.24) | 147 (73.1%) | 1196 | 48.43 (15.26) | 541 (45.2%) |
| Sheba cohort | 140 | 43.64 (7.15) | 86 (61.4%) | 551 | 33.22 (7.84) | 316 (57.4%) |
| Columbia cohort | 158 | 42.96 (6.15) | 103 (65.2%) | 500 | 64.29 (10.02) | 177 (35.5%) |
| Pavlov and Human brain institute cohort | 28 | 43.29 (6.54) | 13 (46.4%) | 417 | 73.61 (10.49) | 172 (41.2%) |
| UKBB | 75 | 46.19 (2.72) | 32 (42.7%) | 1,932 | 56.52 (7.94) | 1,090 |
|  |  |  |  |  |  | (56.4%) |
| AMP-PD | 191 | 46.42 (4.31) | 114 (59.7%) | 3,093 | 69.76 (13.02) | 1,496<br>(48.4%) |
| All cohorts | 793 | - | - | 7,688 | - | - |

### Gene selection, sequencing and processing

We designed a targeted lysosomal gene panel comprising 36 genes, which was applied to four cohorts collected at McGill University. The panel included genes encoding (1) enzymes involved in ceramide, glycosphingolipid, and ganglioside metabolism (including acid ceramidase, hexosaminidases, cathepsins, and components of the ceramide/S1P pathway); (2) mucopolysaccharidosis (MPS)-related enzymes associated with neuronal lysosomal storage phenotypes; and (3) glycosyl-transferases and sialyltransferases that regulate glycosphingolipid composition and lysosomal membrane trafficking. These genes were selected based on a structured literature review of genes most likely to influence lysosomal substrate processing and lipid homeostasis, processes strongly implicated in Parkinson’s disease biology. The complete gene list, protein functions, and known associated conditions as per OMIM are provided in **Supplementary Table S1**. *GBA1* was not included in this analysis because its association with Parkinson’s disease is well established and has been analysed separately in our cohorts ^24^.

In the McGill cohorts, targeted capture was performed using molecular inversion probes (MIPs), following our protocol (https://github.com/gan-orlab/MIP_protocol) ^25^. The libraries were sequenced on Illumina NovaSeq 6000 platform (paired-end 100 bp) at the Genome Quebec Innovation Centre. Sequence reads were aligned to the hg19 using BWA, with base-quality recalibration and variant calling conducted using GATK v3.8, joint genotyping was performed across all sequencing batches and the resulting variants were subsequently mapped from hg19 to hg38 using CrossMap^26^. For external datasets, the same 36 genes were extracted from the UKBB and AMP-PD genome sequencing data (hg38).

### Quality control

Variant- and sample-level quality control (QC) was performed following established pipelines used in our previous sequencing studies, with adaptations for each dataset ^5,6^. In the McGill cohorts, variants were excluded if the call rate was below 90%, genotype quality (GQ) below 20, or sequencing depth below 30X. Multi-allelic sites were normalized, and variants deviating from Hardy–Weinberg equilibrium in controls (*P* < 1×10) were removed. Variants supported by fewer than 25% of reads carrying the alternate allele were also excluded to minimize false positives. At the sample level, we excluded individuals with an average genotyping rate below 90%, discordant genetic and reported sex, heterozygosity outliers, or relatedness (first- or second-degree relatives). In the McGill cohorts, ethnicity was included as a covariate in all association analyses to minimize potential confounding.

For the UKBB and AMP-PD whole-genome datasets, we applied a sequencing depth cut-off of >20x and restricted analyses to participants of European ancestry, determined using principal components or ancestry labels (UKBB field 21000). Population structure was assessed by principal component analysis (PCA), and ancestry outliers were excluded. The AMP-PD variant QC additionally followed the official consortium pipeline (https://amp-pd.org/whole-genome-data).

### Annotation, grouping, and domains

Variants were annotated with ANNOVAR ^27^. Rare variants were defined as minor allele frequency (MAF) ≤ 0.01 within each study cohort. Burden analysis for rare variants was performed in several groups: all rare variants, nonsynonymous variants; loss-of-function (nonsense, frameshift, canonical splice) and predicted missense damaging (CADD ≥ 20). For domain-based burden, variants were mapped to UniProt/Pfam-defined functional regions (catalytic, ligand-binding, luminal/membrane segments) and analyses were repeated using the same variant groupings.

### Statistical analysis

Within each cohort, we tested rare-variant association using SKAT-O (R package) ^28^, adjusting for age, sex and ethnicity when available. We meta-analysed rare variants using MetaSKAT. False discovery rate correction using the Benjamini–Hochberg method was applied. Corrected Pfdr<0.05 was considered significant, uncorrected P<0.05 is reported as nominal.

## Results

### Rare-variant analysis identifies an association of *ST3GAL3* with Parkinson’s disease

Targeted sequencing in McGill cohorts achieved high on-target performance (Average >95% bases at ≥30×), enabling confident rare-variant detection. To access rare variant burden associations across lysosomal genes, we performed SKAT-O analyses in the McGill cohorts, UKBB and AMP-PD cohorts. The full list of coding rare variants across the 36 targeted lysosomal genes in all studied cohorts is provided in **Supplementary Table 2**.

Meta-analyses across the McGill, UKBB, and AMP-PD cohorts identified an association between all rare variants in *ST3GAL3* and Parkinson’s disease (Pfdr=0.04). Several additional genes showed nominal associations (P<0.05), including *HGSNAT, ASAH1, CTSD, HEXA, ST3GAL4* and *SGPP1* (**Table 2; Supplementary Tables S3–S4**). These signals mostly reflected the aggregate contribution of multiple rare alleles, with no single variant consistently driving the gene-level associations across cohorts.

**Table 2.**
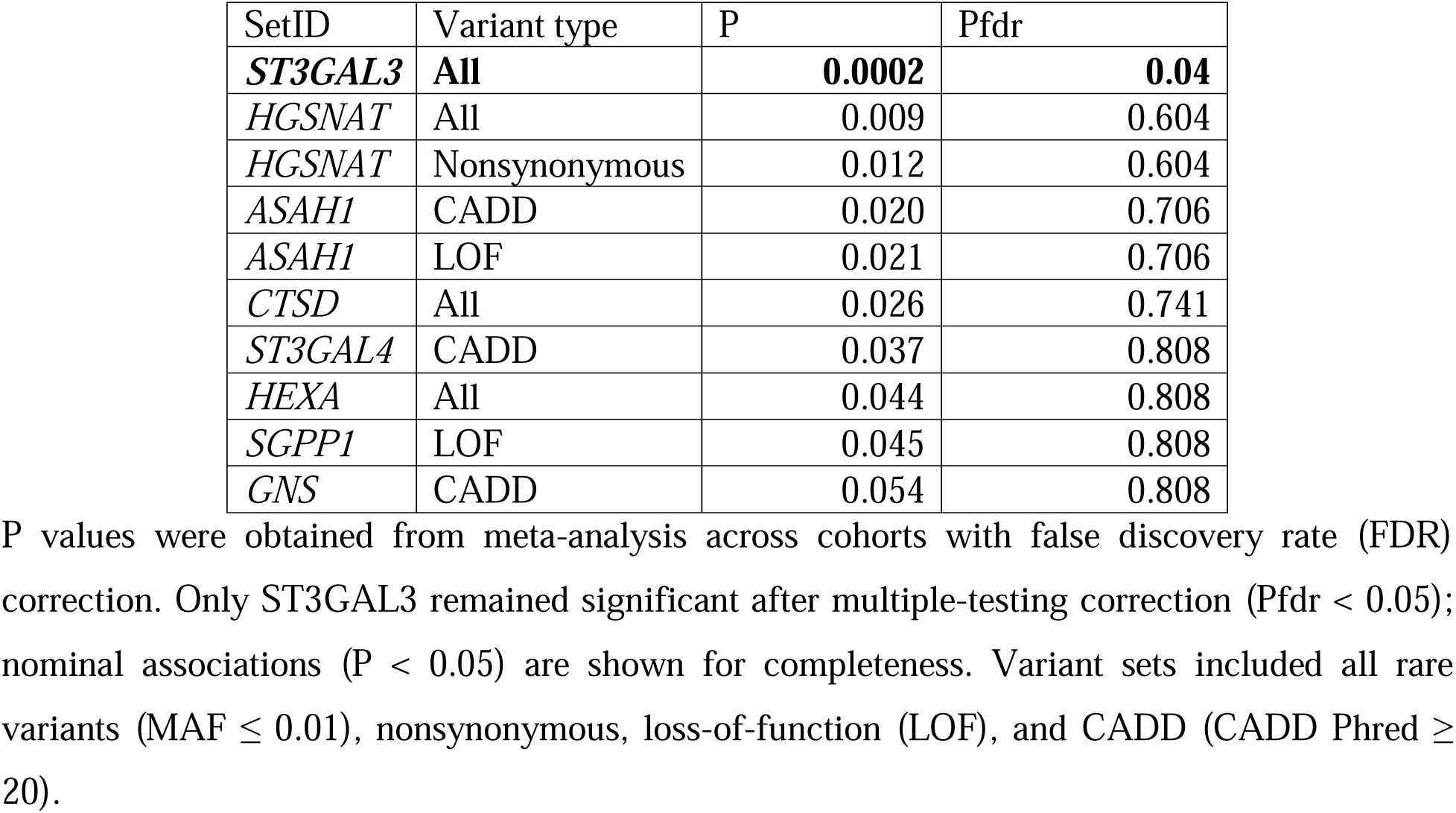
Rare-variant burden meta-analysis associations across lysosomal genes in Parkinson’s disease.

Single-variant analysis highlighted several variants enriched in cases. In *ASAH1*, the nominal gene-level signal was influenced by the missense variants p.V369I, which was observed in a total of 14 Parkinson’s disease cases and no controls across the Pavlov and Human Brain and Columbia cohorts. However, this variant did not demonstrate consistent enrichment among cases across other cohorts. In addition, a rare loss-of-function stop-gain variant in *ASAH1* p.Q95X was observed across three independent cohorts. This variant was detected in 13 Parkinson’s disease cases and 5 controls in the McGill cohort (OR = 2.79, P = 0.052), 23 cases and 6 controls in the Columbia cohort (OR = 1.72, P = 0.239), and 9 cases and 4 controls in the Pavlov and Human Brain cohort (OR = 1.85, P = 0.309). Although none of these associations reached statistical significance individually, the recurrent observation of this loss-of-function variant across cohorts suggests a potential contribution to disease risk. Another recurrent variant was *NAGLU* p.T441M, which was observed in Parkinson’s disease cases across multiple cohorts (total N=12), with low or absent counts (total N=5) in controls across all cohorts but did not reach significance in individual cohorts due to its rarity.

In early-onset Parkinson’s disease, rare-variant burden meta-analysis identified only nominal associations (P<0.05) across several genes, including *GRN, ST3GAL5, UGCG, HEXB, MANBA, PPT1, HGSNAT, CERK, B4GALT1, B4GALT7, ST3GAL3*, and *SGSH*, none of which survived correction for multiple testing (**Supplementary Tables S5-S6**).

### Domain based burden analysis identified significant association in sphingolipid-related genes

Domain-based burden analysis identified a significant enrichment of nonsynonymous variants within the beta-acetyl-hexosaminidase–like domain of *HEXA* (P=8.0*10^-4^), which did not survive correction for multiple testing (Pfdr = 0.15). Additional domain-level associations were nominal (P<0.05) and involved *HGSNAT*, *ASAH1*, *CERK*, and *GRN*. Detailed domain definitions, along with cohort-specific and meta-analysis results, are provided in **Supplementary Tables S7–S8**.

Within *HGSNAT,* the p.L406W variant was enriched in Parkinson’s disease cases in three McGill-sequenced cohorts (Pavlov and Human Brain, Sheba, and Columbia; 8 cases vs 1 control), while showing an opposite direction of effect in the McGill cohort (4 cases vs 13 controls). The variant was not detected in other cohorts after quality control. For *ASAH1*, both variants described above (p.V369I and p.Q95X) map to the ceramidase domain and together account for the observed Ceramidase domain-level association.

### Domain-based burden analysis in early-onset Parkinson’s disease

In early-onset Parkinson’s disease, domain-based analyses identified significant associations in *NAGLU* (Pfdr=7.33*10^-6^), and *ST3GAL5* (Pfdr=0.03) **(Supplementary Tables S9–S10)**. The association in *NAGLU* was primarily driven by the p.Ser141Thr variant, which was observed in seven early-onset Parkinson’s disease cases of Jewish or African/African-American ancestry and in one control of African ancestry. This variant is common in African populations, and additional replication in independent cohorts of Jewish, African, and African-American ancestry is required to confirm this association. Single-variant analyses did not identify other significant associations.

## Discussion

In this study, we evaluated the burden of rare variants across multiple lysosomal genes in Parkinson’s disease using multi-cohort sequencing data. Our findings implicate pathways involved in sialylation and glycosylation (*ST3GAL3, ST3GAL4, ST3GAL5*), ganglioside metabolism (*HEXA*), ceramide and sphingolipid biology (*ASAH1, SGPP1, CERK*), and lysosomal proteolysis (*CTSD*) in Parkinson’s disease susceptibility, further extending the lysosomal genetic architecture beyond the established role of *GBA1*.

Among the genes analysed, *ST3GAL3*, *ST3GAL4*, and *ST3GAL5* showed evidence of rare-variant burden, implicating sialylation pathways in Parkinson’s disease susceptibility. These three sialyltransferases have complementary roles in ganglioside and glycoprotein biosynthesis^29^. *ST3GAL3* encodes ST3Gal-III, a sialyltransferase that regulates terminal sialylation of glycoproteins and glycolipids ^30^, including major brain gangliosides GD1a and GT1b essential for neuronal function ^31^. Biallelic pathogenic variants in *ST3GAL3* are associated with developmental and epileptic encephalopathy-15 (DEE15)^32^. *ST3GAL5* encodes GM3 synthase, catalyzing the first step in ganglioside biosynthesis, and pathogenic variants result in profound neurodevelopmental disorder ^33^. Reduced GM1 and related gangliosides have been documented in Parkinson’s disease substantia nigra ^34^, and GM1 directly modulates alpha-synuclein aggregation ^35^. Consistent with impaired ganglioside metabolism, plasma lipidomic studies have demonstrated elevated GM3 ganglioside concentration in Parkinson’s disease patients compared to controls ^36^. Recent large-scale exome-wide rare-variant burden analyses have implicated B3GNT3 ^37^, an enzyme that synthesizes poly-N-acetyllactosamine chains serving as substrates for ST3GAL-mediated sialylation, providing independent genetic support for perturbation of this pathway in Parkinson’s disease. Together, the concordant signals across *ST3GAL3*, *ST3GAL4*, and *ST3GAL5* support a model in which altered sialylation affects glycosphingolipid composition and associated with lysosomal dysfunction and Parkinson’s disease pathogenesis.

Domain-based analyses provided additional resolution of genetic signal, identifying an enrichment of nonsynonymous variants within the beta-acetyl-hexosaminidase–like domain of *HEXA*. Biallelic pathogenic mutations in HEXA cause Tay-Sachs disease (GM2 gangliosidosis), which ranges from infantile to juvenile and adult/late-onset forms. In late-onset forms, motor neuron disease, cerebellar ataxia, and psychiatric symptoms are common. HEXA plays a central role in lysosomal ganglioside degradation by catalyzing the conversion of GM2 to GM3^38^, functionally opposing the biosynthetic pathway mediated by *ST3GAL5* (Figure 1). Disruption of ganglioside homeostasis has been linked to alpha-synuclein aggregation, impaired lysosomal flux, and neurodegeneration ^39^. Consistent with this model, gangliosides including GM2 and GM3 are elevated across multiple brain regions in *GBA1*-associated Parkinson’s disease, indicating that lysosomal dysfunction propagates upstream through the ganglioside degradation pathway ^40^. Interestingly, increased HEXA activity in rodent models prevents alpha-synuclein lipid associations and protects dopaminergic neurons ^41,42^. In line with impaired lysosomal metabolism, HEX activity is reduced in the substantia nigra in Parkinson’s disease ^41^. Therapeutic strategies targeting ganglioside metabolism, may offer disease-modifying potential across genetic and idiopathic forms of Parkinson’s disease.

**Figure 1.**
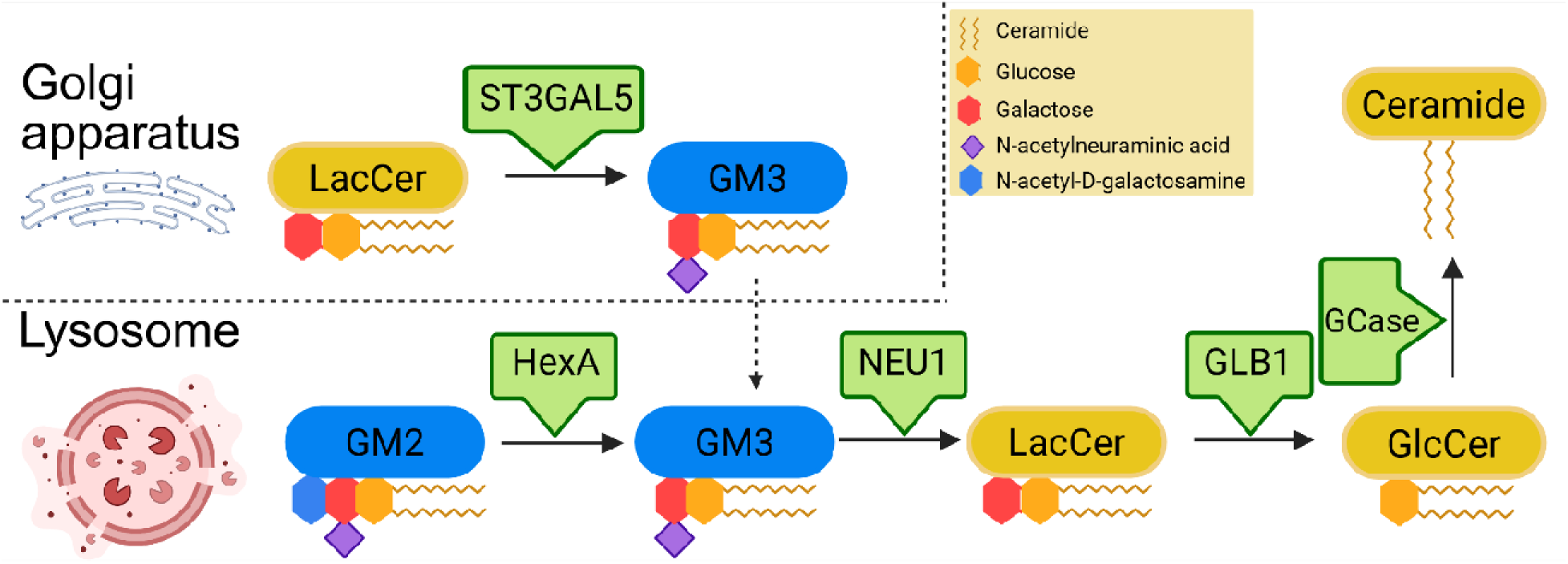
Ganglioside biosynthesis and lysosomal degradation of GM3. Gangliosid biosynthesis is initiated in the late Golgi compartments, where lactosylceramide (LacCer) i converted to GM3 by ST3GAL5 (GM3 synthase). GM3 is transported to the plasma membrane and subsequently delivered to lysosomes via membrane trafficking and endocytosis (dashed arrows). Within lysosomes, ganglioside degradation proceeds in a unidirectional, stepwise manner from more complex to simpler species, including conversion of GM2 to GM3 by β-hexosaminidase A (HEXA, with GM2 activator protein). GM3 is further degraded to LacCer by neuraminidase 1 (NEU1), followed by conversion of LacCer to glucosylceramide (GlcCer) by β-galactosidase (GLB1). GlcCer is subsequently degraded to ceramide by glucocerebrosidase (GCase). This figure was created with BioRender.com.

Additional nominal associations were observed in genes involved in ceramide and sphingosine-1-phosphate handling, including *ASAH1*, *SGPP1*, and *CERK*. Ceramide and sphingolipid balance are key determinants of lysosomal membrane integrity, autophagy initiation, and cellular stress responses ^43^. Disruption of these pathways has been shown to impair alpha-synuclein clearance and exacerbate neurotoxicity in experimental models, supporting the biological relevance of these findings ^43^. *CTSD* is one of the lysosomal proteases and may degrade alpha-synuclein ^44,45^.

Our findings extend previous observations that Parkinson’s disease is characterized by an aggregate lysosomal genetic burden independent of *GBA1* status, while also illustrating the challenges of identifying individual lysosomal genes with modest signal ^8,10–12^. Several genes highlighted here, including *CTSD*, *ASAH1*, and *GRN*, have previously been implicated in Parkinson’s disease or related neurodegenerative phenotypes ^8^, providing additional support to their potential involvement in the disease pathophysiology. However, most associations identified in this study were nominal and did not survive stringent correction for multiple testing. This pattern is consistent with prior rare-variant burden studies in Parkinson’s disease, in which *GBA1* represents a notable outlier with a comparatively large effect, whereas other lysosomal genes appear to confer smaller increments of risk that require substantially larger sample sizes to detect reliably.

It is also notable that several lysosomal genes previously implicated in Parkinson’s disease were not significantly associated in the present analysis. This likely reflects inherent limitations of rare-variant burden testing, which is best powered to detect dominant or semi-dominant effects and may be less sensitive to recessive mechanisms or ultra-rare variants. In addition, pathogenic variants in some lysosomal genes are exceedingly rare or associated with phenotypes that differ from typical late-onset Parkinson’s disease, such as earlier age at onset or distinct clinical features. As the majority of cases included in this study represent the general Parkinson’s disease population with later onset, carriers of highly penetrant recessive mutations may be underrepresented.

Analysis of early-onset Parkinson’s disease identified strong domain-based associations in *NAGLU*, and *ST3GAL5*. These findings warrant cautious interpretation. In particular, the association observed in *NAGLU* was largely driven by a single variant (p.Ser141Thr) that is common in African populations gnomAD (MAF=0.067), underscoring the importance of ancestry-aware analyses in rare-variant studies. Replication is required in larger and more ancestrally diverse sequencing studies to refine effect estimates and identify ancestry-specific risk alleles.

Several limitations should be acknowledged. First, this study integrates data generated using different sequencing platforms and capture strategies, necessitating dataset-specific quality control thresholds that may influence variant detection sensitivity. Second, although analyses were adjusted for available covariates, residual population stratification cannot be fully excluded, particularly in rare-variant analyses and domain-based tests. Third, kernel-based burden tests do not provide information on the directionality of individual variants, limiting interpretation of whether specific alleles confer risk or protection. Fourth, our analysis focused on single-nucleotide variants and small indels and did not include copy number variants. Finally, functional validation was beyond the scope of this study and the biological consequences of the identified genes remain to be established.

In conclusion, these data provide further genetic evidence that rare variants across multiple lysosomal pathways contribute to Parkinson’s disease susceptibility, implicating sialylation, ganglioside metabolism, ceramide biology, and lysosomal proteolysis. The convergence of genetic associations in both biosynthetic (*ST3GAL3, ST3GAL4, ST3GAL5*) and degradative (*HEXA*) arms of the ganglioside pathway is particularly compelling. While *GBA1* remains the strongest lysosomal risk gene, several additional lysosomal genes previously implicated in smaller rare-variant burden studies ^8^ remained nominally associated in the present analysis despite a substantial increase in sample size. Although these signals did not survive correction for multiple testing, their consistency across studies suggests reproducible but modest effects. This pattern likely reflects both the small effect sizes of non-*GBA1* lysosomal genes and methodological constraints of kernel-based burden tests that aggregate variants by class. Larger, ancestrally diverse cohorts and complementary functional studies will be required to refine effect estimates and clarify the contribution of these genes to Parkinson’s disease pathogenesis.

## Data availability

All supplementary materials, including additional tables and figures, are available in the Supplementary Materials section. The McGill cohorts are partially available through The Canadian Open Parkinson Network (C-OPN). Access, including genetic data, can be requested through the C-OPN data access committee (https://copn-rpco.ca/submit-a-request/). The AMP-PD data was assessed using the Terra platform, https://amp-pd.org/. The UKBB was acquired using WGS data from the UK Biobank Research Analysis Platform (https://www.ukbiobank.ac.uk/) under application number 45551.

## Acknowledgements

We would like to thank the participants in the different cohorts for contributing to this study. S.C.P. is supported by a Canadian Institutes of Health Research (CIHR) Canada Graduate Scholarship – Doctoral (CGS-D) award. Z.G.O. is supported by the Fonds de recherche du Québec—Santé (FRQS) Chercheurs-boursiers award, in collaboration with Parkinson Quebec, and is a William Dawson Scholar. The access to part of the participants for this research has been made possible thanks to the Quebec Parkinson’s Network (http://rpq-qpn.ca/en/).

Data used in the preparation of this article were obtained from the Accelerating Medicine Partnership® (AMP®) Parkinson’s Disease (AMP PD) Knowledge Platform. For up-to-date information on the study, visit https://www.amp-pd.org. Data for this article were also obtained in January 2023 from the AMP-PD Knowledge Platform as per release 2.5. Since this release more recent versions have been made available. The AMP® PD program is a public-private partnership managed by the Foundation for the National Institutes of Health and funded by the National Institute of Neurological Disorders and Stroke (NINDS) in partnership with the Aligning Science Across Parkinson’s (ASAP) initiative; Celgene Corporation, a subsidiary of Bristol-Myers Squibb Company; GlaxoSmithKline plc (GSK); The Michael J. Fox Foundation for Parkinson’s Research ; Pfizer Inc.; AbbVie Inc.; Sanofi US Services Inc.; and Verily Life Sciences. ACCELERATING MEDICINES PARTNERSHIP and AMP are registered service marks of the U.S. Department of Health and Human Services. Clinical data and biosamples used in preparation of this article were obtained from the (i) Michael J. Fox Foundation for Parkinson’s Research (MJFF) and National Institutes of Neurological Disorders and Stroke (NINDS) BioFIND study, (ii) Harvard Biomarkers Study (HBS) and the Stephen & Denise

Adams Center for Parkinson’s Disease Research of Yale School of Medicine (CPDR-Y), (iii) National Institute on Aging (NIA) International Lewy Body Dementia Genetics Consortium Genome Sequencing in Lewy Body Dementia Case-control Cohort (LBD), (iv) MJFF LRRK2 Cohort Consortium (LCC), (v) NINDS Parkinson’s Disease Biomarkers Program (PDBP), (vi) MJFF Parkinson’s Progression Markers Initiative (PPMI), and (vii) NINDS Study of Isradipine as a Disease-modifying Agent in Subjects With Early Parkinson Disease, Phase 3 (STEADY-PD3) and (viii) the NINDS Study of Urate Elevation in Parkinson’s Disease, Phase 3 (SURE-PD3). BioFIND is sponsored by The Michael J. Fox Foundation for Parkinson’s Research (MJFF) with support from the National Institute for Neurological Disorders and Stroke (NINDS). The BioFIND Investigators have not participated in reviewing the data analysis or content of the manuscript. For up-to-date information on the study, visit michaeljfox.org/news/biofind. Genome sequence data for the Lewy body dementia case-control cohort were generated at the Intramural Research Program of the U.S. National Institutes of Health. The study was supported in part by the National Institute on Aging (program #: 1ZIAAG000935) and the National Institute of Neurological Disorders and Stroke (program #: 1ZIANS003154). The Harvard Biomarker Study (HBS) is a collaboration of HBS investigators [full list of HBS investigators found at https://www.bwhparkinsoncenter.org/biobank/ and funded through philanthropy and NIH and Non-NIH funding sources. The Stephen & Denise Adams Center for Parkinson’s Disease Research of Yale School of Medicine is funded through philanthropy and NIH and non-NIH funding sources. The HBS and CPDR-Y Investigators have not participated in reviewing the data analysis or content of the manuscript. Data used in preparation of this article were obtained from The Michael J. Fox Foundation sponsored LRRK2 Cohort Consortium (LCC). The LCC Investigators have not participated in reviewing the data analysis or content of the manuscript. For up-to-date information on the study, visit https://www.michaeljfox.org/biospecimens). PPMI is sponsored by The Michael J. Fox Foundation for Parkinson’s Research and supported by a consortium of scientific partners: [list the full names of all of the PPMI funding partners found at https://www.ppmi-info.org/about-ppmi/who-we-are/study-sponsors]. The PPMI investigators have not participated in reviewing the data analysis or content of the manuscript. For up-to-date information on the study, visit www.ppmi-info.org. The Parkinson’s Disease Biomarker Program (PDBP) consortium is supported by the National Institute of Neurological Disorders and Stroke (NINDS) at the National Institutes of Health. A full list of PDBP investigators can be found at https://pdbp.ninds.nih.gov/policy. The PDBP investigators have not participated in reviewing the data analysis or content of the manuscript. The Study of Isradipine as a Disease-modifying Agent in Subjects With Early Parkinson Disease, Phase 3 (STEADY-PD3) is funded by the National Institute of Neurological Disorders and Stroke (NINDS) at the National Institutes of Health with support from The Michael J. Fox Foundation and the Parkinson Study Group. For additional study information, visit https://clinicaltrials.gov/ct2/show/study/NCT02168842. The STEADY-PD3 investigators have not participated in reviewing the data analysis or content of the manuscript. The Study of Urate Elevation in Parkinson’s Disease, Phase 3 (SURE-PD3) is funded by the National Institute of Neurological Disorders and Stroke (NINDS) at the National Institutes of Health with support from The Michael J. Fox Foundation and the Parkinson Study Group. For additional study information, visit https://clinicaltrials.gov/ct2/show/NCT02642393. The SURE-PD3 investigators have not participated in reviewing the data analysis or content of the manuscript. Figure 1 was created with BioRender.com.

